# Identification of COVID-19 Vaccine-Hesitancy Predictors in the United States

**DOI:** 10.1101/2023.07.17.23292772

**Authors:** Enrique M. Saldarriaga

## Abstract

Vaccine hesitancy is complex, multi-causative phenomenon that undermines public health efforts to contain the spread of infectious diseases. Improving our understanding of the drivers of vaccine hesitancy might improve our capacity to address it. We used the results of the May 2021 ASPE’s survey on COVID-19 vaccine-hesitancy which estimated the proportion of adults that felt hesitant of unsure towards taking the COVID-19 vaccine when it becomes available at the county-level. We developed a prediction model to identify the most important predictors of vaccine-hesitancy. The potential predictors included demographic characteristics, the CDC’s social vulnerability index, and the Republican Party’s voting share in the 2020 presidential election as a proxy of political affiliation, both at the county-level. The most important drivers of hesitancy included low educational attainment, proportion of Black/African American population, and political affiliation. These results deepen our understanding of the phenomenon and could help design more targeted interventions to reduce hesitancy in specific sub-groups of the population.

## Introduction

The COVID-19 pandemic has caused over 6 million hospitalizations and over 1 million deaths in the United States.[1] The COVID-19 vaccine was developed in record time[2] and rapidly became the most effective measure to control the spread of the virus and mitigate the severity of COVID-19 illness.[3] Despite the efforts of federal agencies, namely the Food and Drug Administrations (FDA) and the Centers for Disease Control and Prevention (CDC), to communicate to the public the proven safety and efficacy of the vaccines,[4] vaccine hesitancy has emerged as a major challenge in achieving global vaccination coverage and controlling the pandemic.[5] Thus, the efforts of local health departments across the country, fell short in administering vaccines.

Vaccine hesitancy refers to the delay in acceptance or refusal of vaccination despite the availability of vaccination services. This is a complex, multicausal phenomena that has arisen as a major public health concern during the last decade.[6] The reasons for vaccine hesitancy include factors related to individual beliefs, attitudes, and behaviors, as well as contextual factors such as social, cultural, and political factors.[7] Socioeconomic and demographic factors have been identified as important predictors of vaccine hesitancy. Studies have shown that individuals with lower education, income, and social status are more likely to be vaccine hesitant.[8,9] Other demographic factors such as age, gender, and race/ethnicity have also been found to be associated with vaccine hesitancy. However, the relationship between socioeconomic and demographic factors and vaccine hesitancy may vary across different populations and contexts.[8,10] In the specific context of the COVID-19 pandemic, misinformation and conspiracy beliefs had played an important role in creating vaccine hesitancy.[11]

Understanding the predictors of vaccine hesitancy is crucial for designing future vaccination campaigns and policies to effectively overcome vaccine hesitancy and increase vaccination coverage. Especially because the societal polarization around the relevance and safety of public health activities during the COVID-19 pandemic, might have spilled over to other diseases.[12] The goal of this study is to identify the most important predictors of COVID-19 vaccine hesitancy in the United States from an ecological perspective and employing machine learning methods to increase the accuracy of our predictions. The findings of this study can help identify the population groups most likely to be reluctant to accept the vaccine and therefore help in the designing of targeted interventions and policies to promote vaccination coverage.

## Methods

### Study Population

A cross-sectional survey was conducted between May 26 and June 7, 2021, as part of the U.S. Census’ Bureau Household Pulse Survey (HPS) to measure levels of hesitancy towards the COVID-19 vaccine.[13] The survey had a multiple-choice question “Once a vaccine to prevent COVID-19 is available to you, would you get a vaccine?”, with five possible answers: “definitely get a vaccine”; “probably get a vaccine”; “unsure”; “probably not get a vaccine”; “definitely not get a vaccine.” A the time of the survey, the vaccine was under development and would not be approved until August 23, 2021.[14]

The U.S. Department of Health and Human Services’ Office of the Assistant Secretary for Planning and Evaluation (ASPE) used the household-level survey results, to create estimates at the county-level of the proportion of people within a county that fell into each of the mutually-exclusive categories of hesitancy.[15] For the analysis, we classify the proportion that felt strongly against taking the vaccine as Strongly Hesitant; and the proportion that answered “unsure” and “probably not get a vaccine” as Hesitant or Unsure. Thus, we obtained two metrics of vaccine hesitancy intensity, for all 3,142 counties in the U.S. and the District of Columbia.

### Measurement of demographic information

We used the American Community Survey 5-years to obtain county-level demographic information.[16] We selected variables with observed association with vaccine hesitancy in the literature[10,11,17]. The final prediction set included the following categories: race and ethnicity, marital status, age, biological sex, poverty level respect to federal poverty line, income, internal immigration (people who moved to the county in the las 12 months from within the US), foreign immigration (people who moved to the county in the las 12 months from outside the US), schooling, urbanicity, employment, and the county’s GINI coefficient as a measure of inequality.

In addition, we included the CDC’s Social Vulnerability Index, a relative measure of an areas’ risk to a hazardous event of any kind.[18]

### Political Affiliation

Misinformation plays an important role in the generation of vaccine hesitant feelings.[19,20] It has been documented that people whose political preferences lean towards the Republican Party in the United States were more prone to consume wrongful and misleading information about the safety, efficacy, and importance of the COVD-19 Vaccine.[21–23] Using a web-based survey, Sargent, et al found that Democratic-leaning people were 2.4 (95% confidence interval 2.2., 2.7) times more likely to be vaccinated and 1.8 (95%CI 1.5, 2.2.) time more likely to be receptive towards vaccination, than their Republican-leaning peers.[24] To account for the differences in vaccine hesitancy associated with political affiliation, we included the county-level proportion of votes that the Republican Party obtained in the 2020 Presidential Election. We obtained the data from the MIT Sloan School of Management’s repository.[25]

### Statistical Analysis

We followed a machine learning algorithm to identify the most important predictors of vaccine hesitancy expressed in two outcomes, Hesitant or Unsure and Strongly Hesitant. We used least absolute shrinkage and selection operator (LASSO) regression to identify the most important predictors for each outcome. LASSO is type of regularized regression, that introduces a penalty in the estimation of a variable’s coefficient for collinearity and low explanatory power over the dependent variable.[26] This approach allows us to introduce variables that are seemingly correlated (e.g., income, schooling) in the predictor set, and let the model identify their explanatory capacity conditional to all variables included.

The final data set included only counties with complete information for all potential predictors. We split the sample into a training set equivalent to 80% of the observation, randomly selected, and a test set, equivalent to 20%. Thus, we can assess the prediction accuracy of the model out-of-sample. To train the model, we used leave-one-out cross validation to estimate the optimum lambda, with the Mean Absolute Error (MAE) as the loss function to minimize. We used the coefficients from the best model to predict in the test set. We report the MAE in the test set.

We show the importance of the demographic characteristics in explaining vaccine hesitancy in two ways. First, we used exact post-selection inference to estimate confidence intervals via the sandwich standard error estimation, to show the level of uncertainty associated with a variable’s coefficient.[27,28] This information indicates whether changes in a demographic variable are expected to be associated to changes in the outcome. Second, we plot the values of the coefficients at every value of the log of the LASSO penalty term, the lambda. The highest the penalization, the fewer variables will enter the model, remaining only those with the highest explicative power.[29] Thus, this plot provides a visual representation of which variables are included at every level of penalization, allowing for a qualitative description of the variables’ importance. This plot shows the variable capacity to explain the variability in the outcome, despite the direction of its association.

All analysis were conducted separately for both vaccine hesitancy outcomes, Hesitant or Unsure and Strongly Hesitant. All data management and analyses were conducted in R software.

## Results

Out of 3,142 counties, 2,489 had complete information for all 32 potential predictors identified. Across counties, the proportion of people that reported feeling hesitant or unsure to take the COVID-19 vaccine ranged from 4.9 to 32.3%, with an average of 18.8%. The proportion of people that felt strongly hesitant to take the vaccine varied from 1.9 to 18.2%, with a mean of 8.4%. Both outcomes had a multimodal distribution, where Strongly Hesitant has a greater concentration and a lower range, than Hesitant or Unsure.(Figure 1)

**Figure 1.**
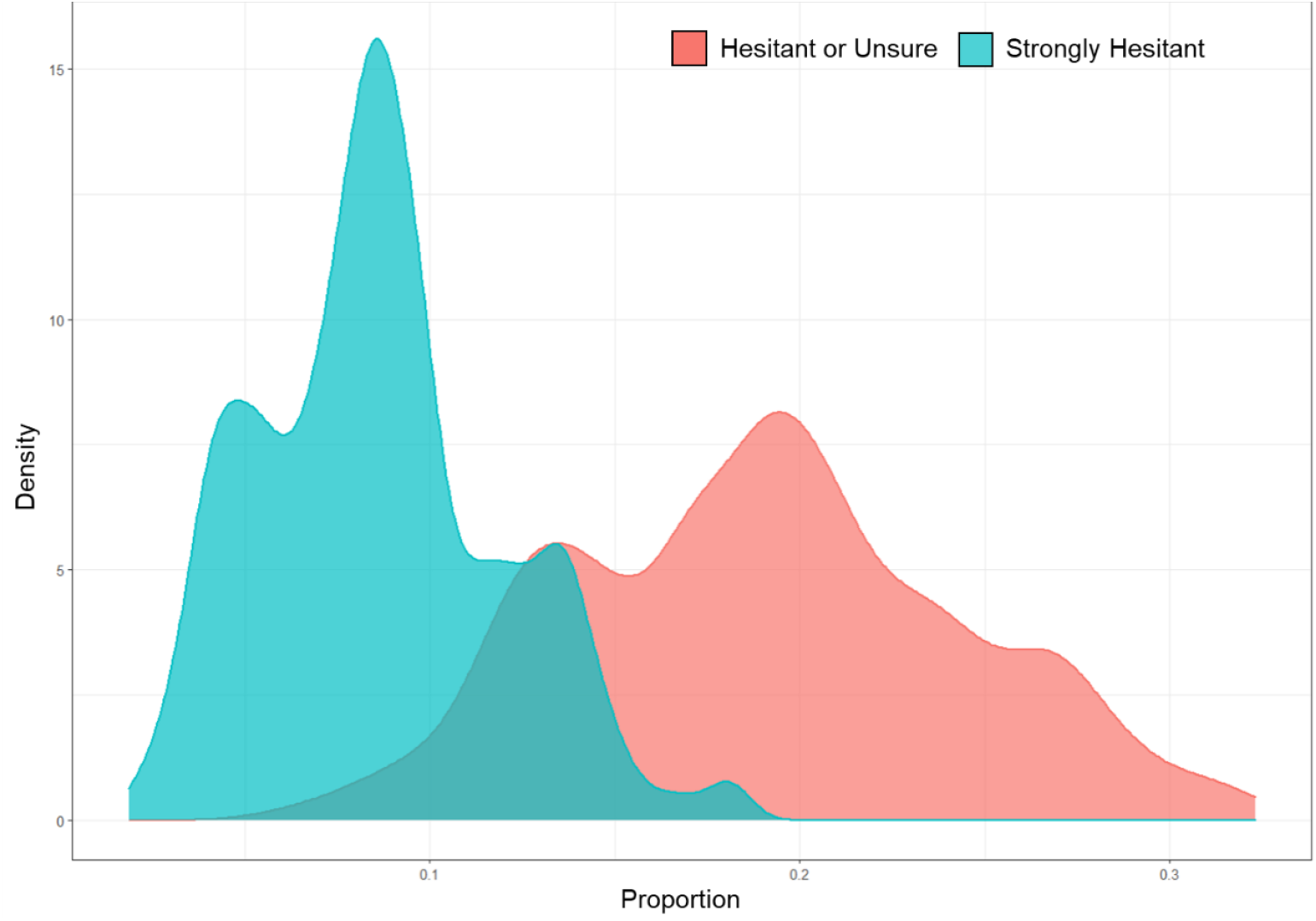
Density distribution of the proportion of people feeling Hesitant or Unsure and Strongly Hesitant to take the COVID-19 vaccine at the county-level.

The models showed good prediction accuracy for both hesitancy outcomes. For the Hesitant or Unsure outcome, we found a MAE of 2.8% in the out-of-sample validation set. This means that our model had an average error of 2.8% when comparing the observed versus the predicted proportion of people hesitant of unsure. At the optimum penalization (i.e., the lambda that minimizes the in-sample MAE), two variables were excluded from the model due to low explanatory power, proportion of people with some college and proportion of unemployed males. We found that an increase of 1% in the proportion of divorcees in a county is associated with a 0.139% (95% confidence interval 0.292, 0.147) increase in the proportion of people that reported feeling hesitant or unsure to take the COVID-19 vaccine.(Table 1, column A) Other variables associated with an increase in hesitancy were proportions of Black/African-American, other race/ethnicity, people with an annual income between 10-25 thousand, people below the federal-level poverty line, internal immigrants, people with a Bachelor’s degree, unemployed females, and uninsured population, higher levels of inequality expressed in the GINI coefficient, and a higher vote-share for the Republican Party in the 2020 Presidential Election. On the other hand, an increase in the proportion of Non-Hispanic Asian population was associated with a 0.349% (95%CI -0.462, -0.250) reduction in the proportion of people that reported feeling Hesitant or Unsure to take the COVID-19 vaccine. Other variables associated with a decrease in hesitancy are the proportion of widowers, of people of Hispanic ethnicity, of people aged 65 years and over, of people living outside the city, and higher levels of SVI.(Table 1, column A).

**Table 1.** Coefficients from the LASSO regression at the optimum level of penalization for the Hesitant or Unsure and Strongly Hesitant outcomes.

|  | (A) Hesitant or Unsure<br>Coefficient (% , 95% CI) | (B) Strongly Hesitant<br>Coefficient (% , 95% CI) |
| --- | --- | --- |
| **Optimum Penalization (Lambda)** | 0.0001004471 | 0.0001229605 |
| **Variables** |  |  |
| Intercept | -0.061 | -0.055 |
| Vote share for the Republican Party | 0.116 (0.1, 0.133)** | 0.064 (0.059, 0.132)** |
| Race/Ethnicity |  |  |
| Non-Hispanic White | Not included | Not included |
| Hispanic | -0.053 (-0.068, -0.039)** | -0.043 (-0.068, -0.039)** |
| Non-Hispanic African American | 0.137 (0.117, 0.157)** | 0.039 (0.016, 0.156)** |
| Non-Hispanic Asian | -0.349 (-0.462, -0.25)** | -0.218 (-0.461, -0.2)** |
| Other | 0.139 (0.105, 0.187)** | 0.1 (0.095, 0.186)** |
| Marital Status |  |  |
| Never Married | Not included | Not included |
| Married | -0.012 (-0.053, 0.021) | 0.011 (-0.053, 0.019) |
| Widowed | -0.276 (-0.413, -0.147)** | -0.171 (-0.41, -0.144)** |
| Divorced | 0.371 (0.292, 0.451)** | 0.224 (0.21, 0.449)** |
| Age |  |  |
| 50 to 54 | Not included | Not included |
| under 18 | 0.067 (-0.001, 0.166) | 0.027 (0, 0.163) |
| 18 to 25 | -0.032 (-0.113, 0.037) | -0.053 (-0.112, 0.031) |
| 25 to 39 | 0.038 (-0.046, 0.154) | 0.015 (-0.038, 0.151) |
| 55 to 64 | 0.123 (-0.007, 0.316) | 0.088 (-0.002, 0.306) |
| Over 65 | -0.305 (-0.386, -0.219)** | -0.172 (-0.381, -0.16)** |
| Sex |  |  |
| Female | Not included | Not included |
| Male | 0.086 (-0.014, 0.198) | 0.039 (-0.011, 0.193) |
| Poverty, respect to federal threshold |  |  |
| Above 150% | Not included | Not included |
| 100-140% | 0.046 (-0.017, 0.124) | 0.02 (-0.014, 0.123) |
| Below 100% | 0.103 (0.042, 0.175)** | 0.042 (0.04, 0.175)** |
| Annual Income |  |  |
| Less10K | Not included | Not included |
| 10-25K | 0.145 (0.044, 0.262)** | 0.065 (0.042, 0.256)** |
| 25-50K | 0.016 (-0.064, 0.149) | OMITTED |
| 50-65K | -0.155 (-0.31, 0) | -0.085 (-0.31, 0) |
| 65+K | -0.134 (-0.226, 0) | -0.06 (-0.224, -0.001)** |
| Internal Immigrants | 0.176 (0.063, 0.313)** | 0.172 (0.061, 0.312)** |
| Foreign Immigrants | -0.124 (-0.416, 0.111) | -0.079 (-0.412, 0.104) |
| Schooling |  |  |
| Less than High School | Not included | Not included |
| High School degree | 0.028 (-0.011, 0.075) | OMITTED |
| Some College | OMITTED | -0.018 (-0.047, 0.033) |
| Bachelor's degree | 0.139 (0.055, 0.225)** | 0.112 (0.052, 0.221)** |
| Graduate Degree | -0.013 (-0.116, 0.043) | -0.027 (-0.113, 0.034) |
| Living outside city | -0.015 (-0.023, -0.006)** | -0.009 (-0.023, -0.005)** |
| Unemployed Male | OMITTED | OMITTED |
| Unemployed Female | 0.069 (0.034, 0.107)** | 0.048 (0.033, 0.105)** |
| Uninsured | 0.05 (0.001, 0.099)** | 0.045 (0.001, 0.098)** |
| GINI Coefficient of Inequality | 0.117 (0.06, 0.177)** | 0.079 (0.059, 0.176)** |
| Social Vulnerability Index (SVI) | -0.025 (-0.037, -0.014)** | -0.009 (-0.037, -0.002)** |

*All variables are expressed in proportions of people with a given characteristic. Therefore, the coefficients represent the percentual increase or decrease in the outcomes associated with a 1% increase in the variables. All coefficients and confidence interval values are rounded to the third decimal point.*
*\*\*Indicates variables whose confidence intervals exclude the reference value of zero and are statistically significant at 95% of confidence.*
*Key. Not included indicates variables not included in the analysis to avoid perfect collinearity because these are the complement of the included variables. For example, "female" is not included because for any given county, this is equal to 100%-"male." Omitted: Variables excluded from the analysis at the optimum penalization for lack of explanatory power over the outcome(s).*

From the coefficients versus log of lambda (i.e., penalty level) plot, we observe that the most important predictor of the Hesitant or Unsure outcome is income, because it has enough explanatory power over the outcome to generate a non-zero coefficient even at the highest penalty level.(Figure 2) As the penalization becomes less stringent, other variables enter the model. The remaining top ten variables include, ranked from highest to lowest: marital status, poverty, employment, schooling, race/ethnicity, political affiliation, age, and health insurance coverage.

**Figure 2.**
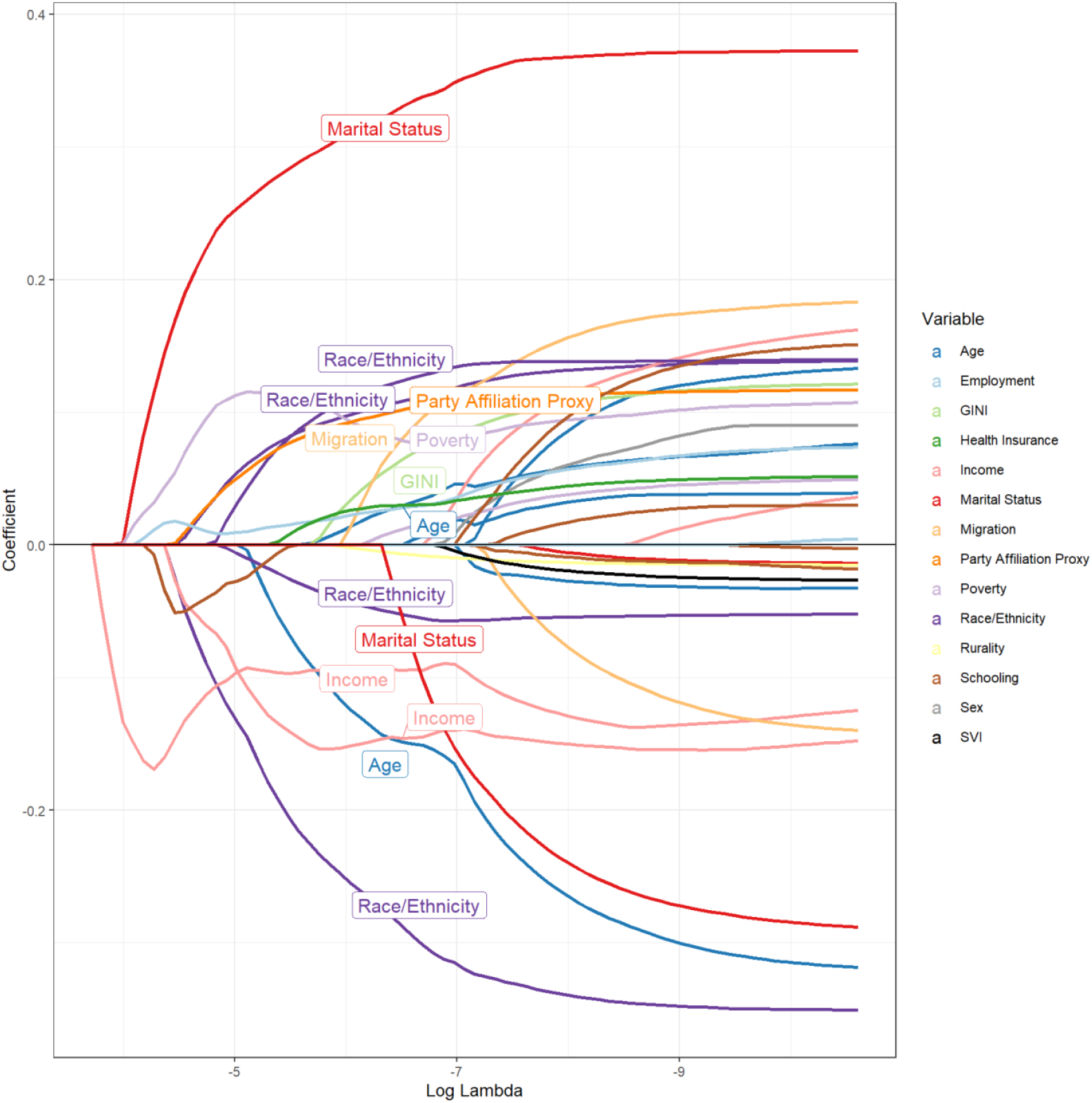
Hesitant or Unsure outcome. Trajectory of the coefficients, color-coded and labeled by variable, at different levels of penalization from the LASSO regression. *Lines represent the coefficients of a specific metric (e.g., Non-Hispanic African American) while color and label represent variable (e.g., Race/Ethnicity)*.

For the Strongly Hesitant outcome, we found a MAE of 2.2% in the out-of-sample validation set. At the optimum penalization, three variables were excluded from the model, proportion of people with an annual income between 25 and 50 thousand dollars, with a high school degree, and proportion of unemployed males. We found that an increase of 1% in the proportion of divorcees in a county is associated with a 0.224% (95% confidence interval 0.21, 0.449) increase in the proportion of people that reported feeling strongly hesitant to take the COVID-19 vaccine.(Table 1, column B) Other variables associated with an increase in strong hesitancy were proportion of Black/African-American, of other race/ethnicity, of people living below the federal poverty line, of having an annual income between 10-25 thousand, of internal immigrants, of having a Bachelor’s degree, of unemployed females, of uninsured, higher levels of economic inequality measured by the GINI coefficient, and a higher share of votes casted for the Republican Party in the 2020 Presidential Election. On the other hand, an increase in the proportion of Non-White Asian population was associated with a 0.218% (95%CI -0.461, -0.2) reduction in the proportion of people that reported feeling Strongly Hesitant to take the COVID-19 vaccine. Other variables associated with a decrease in hesitancy are the proportion of Hispanics, of widowers, of people aged 65 years and over, of people with an annual income of 65 thousand dollars or higher, of people living outside the city, and higher levels of SVI.(Table 1, column B).

From the coefficients versus log of lambda plot, we observe that the most important predictors of the Strongly Hesitant outcome are income and marital status, which have non-zero coefficients at the highest penalty level.(Figure 3) The following eight most important predictors include, in order of highest to lowest: employment, party affiliation, poverty, race/ethnicity, health insurance, migration, and age.

**Figure 3.**
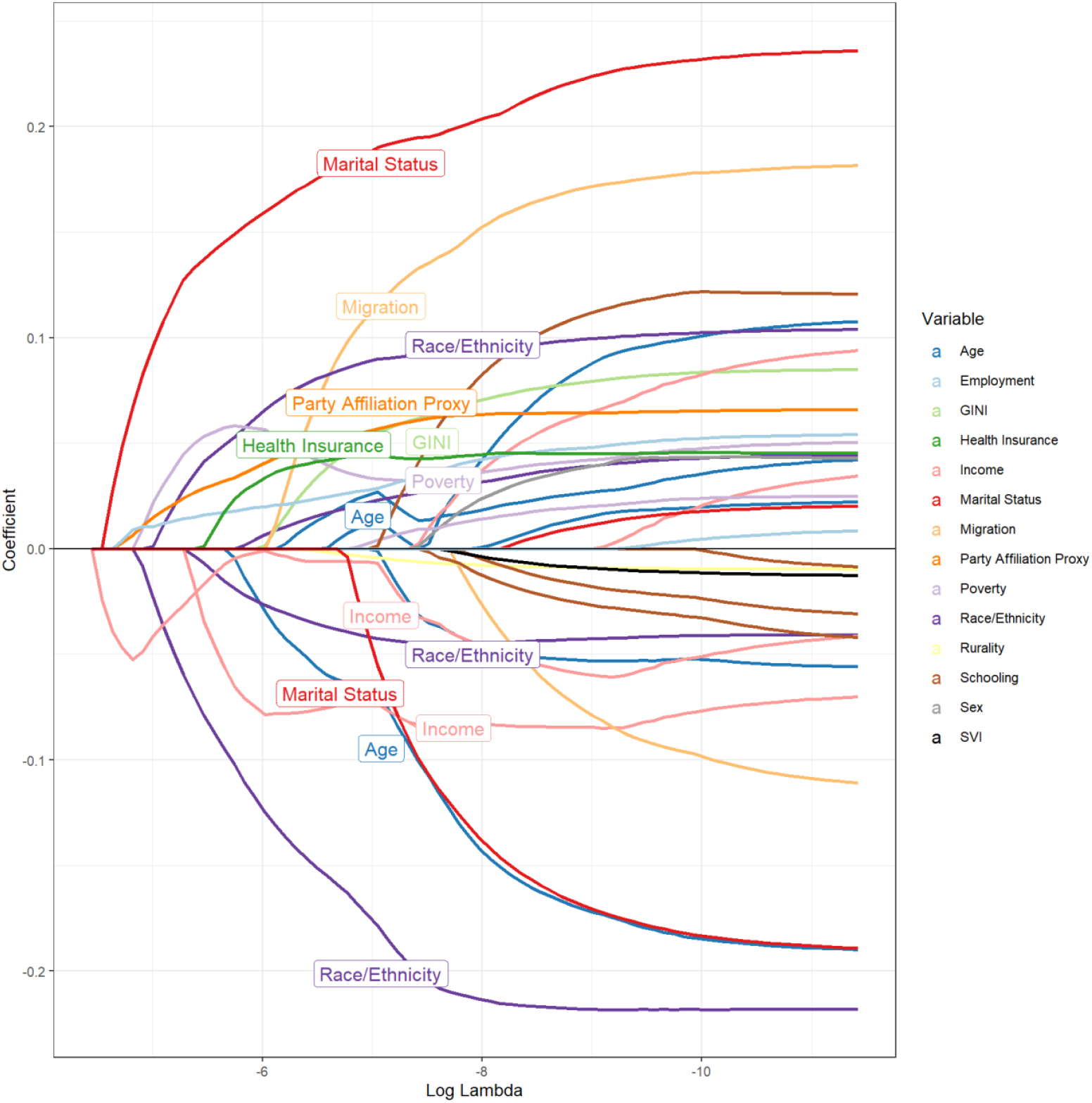
Strongly Hesitant outcome. Trajectory of the coefficients, color-coded and labeled by variable, at different levels of penalization from the LASSO regression. *Lines represent the coefficients of a specific metric (e.g., Non-Hispanic African American) while color and label represent variable (e.g., Race/Ethnicity)*.

## Discussion

We sought to identify the most important predictors of COVID-19 vaccine hesitancy in the United States from a population-level perspective. Hesitancy was measured in two intensities, Hesitant or Unsure, and Strongly Hesitant.[15] We selected potential predictors based on literature review and inclusive of socioeconomic, demographic, and political characteristics. We trained a statistical model on a random selection of 80% of the 2,489 counties with complete information, using a LASSO regression whose penalty parameter was tuned via leave-one-out cross validations. We validated the prediction accuracy of our models in the remaining 20% of the sample. We found that the model had a MEA of 2.82% for the Hesitant or Unsure outcome, and 2.21% for the Strongly Hesitant. The slight improvement in performance observed in the latter outcome might be due to a smoother distribution that more closely resembles a normal distribution than the Hesitant or Unsure counterpart (see Figure 1).

We found great similitude in the characteristics associated to increases and decrements in vaccine hesitancy across both intensities studied. Greater proportions of people living in poverty, with lower income, and unemployment were associated with higher proportion of people feeling Hesitant or Unsure and Strongly Hesitant towards the COVID-19 vaccines. These results are consistent with previous studies that found associated between lower-income individuals and greater vaccine hesitancy.[11,30] One interesting finding was that higher levels of economic inequality, measured by the GINI coefficient, are associated with higher levels of vaccine hesitancy. While we found no evidence of previous studies reporting a direct associated between this metric and hesitancy, the presence of greater economic disparities is consistent with higher proportions of people living under lower economic conditions, which has been documented as a risk factor for vaccine hesitancy.[6]

Regarding race and ethnicity, we found a clear divide between the effects associated with Black/African Americans on one hand, and non-Hispanic Asian and Hispanic on the other one. We found that increases in the proportion of Back/African Americans was associated with higher levels of hesitancy. The distrust of the African American community towards vaccination has been well documented across sub-groups and therapeutic areas, making it a deeply-rooted phenomenon in the US.[31–33] On the other hand, we found that increases in the proportion of Non-Hispanic Asian and Hispanic populations are associated with reductions in vaccine hesitancy. Previous studies have found that Asians in the US had higher levels of trust in the healthcare system than their racial/ethnic minorities counterparts.[33] However, the literature has mixed evidence regarding the level of trust among the Hispanic population. With one study finding higher level of vaccine hesitancy among Hispanic healthcare workers, compared to non-Hispanics Whites[34], while another one found no association after accounting for sociodemographic characteristics such as education and income[35], and several other that suggest that this population is very diverse to find effects above and beyond their educational and economical variability.[31,36,37] Our results suggest that after accounting for socio-economic determinants (including schooling and in employment), higher proportions of Hispanics in a county are associated with lower levels of hesitancy. However, these decrements are much lower than the observed for Asians and African Americans, in absolute terms.

We found no significant associations with sex in either of the hesitancy outcomes. However, we did find a direct effect associated with unemployment among females. This is consistent with previous studies that found that hesitancy is more prevalent among females than males.[30] Regarding marital status there was a clear divide. Higher levels of divorced people were associated with higher levels of hesitancy, and the opposite for higher levels of widowers. Previous studies have found that single people are more likely to be hesitant[30] and on the contrary, married people tend to be less hesitant[35]. Studies have shown that people directly impacted by a disease tend to be less reluctant to take preventive measures to protect their health.[35,38] This could be the case of widowers in our study, especially considering that the ASPE survey took place in May 2021, after half-million of people have died in the United States.[1] Further, this could explain why our results indicate that a higher proportion of people aged 65 years and above, is associated with lower levels of vaccine hesitancy. We did not find any other significant association with age. We found that higher levels of SVI, the CDC’s metric of relative community stress, was associated with lower hesitancy, likely because they may have already experienced a greater burden of COVID-19 or other hazardous events.

Political affinity to the Republican Party, measured as the vote-share of the party in the 2020 Presidential Elections, was associated to higher levels of vaccine hesitancy. One of the most important dimensions in the frameworks that explain the existence and persistence of distrust in vaccines and, more broadly, the healthcare system, is the consumption and believe of misinformation.[39] Further, previous studies have shown that people with low health literacy, prior beliefs regarding the authenticity of the COVID-19 pandemic as a health emergency, and conspiracy beliefs, are important predictors of COVID-19 vaccine hesitancy.[11,30,40] It has been documented how the information sources most frequently consulted and trusted by individuals with affinity for the Republican Party, contained untruthful and misleading information about the importance of the COVID-19 vaccine, its safety and efficacy.[41,42] Unsurprisingly, previous studies have linked a higher affinity to the Republican Party with higher levels of vaccine hesitancy at an individual[38] and ecological[43] levels. In this study we found that party affiliation was an important predictor of vaccine hesitancy in both outcomes, but while it ranked eighth for the Hesitant or Unsure, it ranked fourth for the Strongly Hesitant. Thus, the explanatory power of party affiliation increases as the intensity of the hesitancy does.

These results can be leveraged to increase our understanding of the drivers of hesitancy but even more importantly, to create more detailed profiles of population groups at a higher risk of rejecting vaccine and other preventive measures. The effectiveness of vaccination programs depends on the healthcare system’s capacity to understand the concerns of the population, adequately communicate the importance of these public health actions, and design activities tailored to overcome issues of trust and health literacy.[17] The development of the COVID19 vaccine required a great deal of global resources and political commitment.[2] However, the success of the programs depends on the specific interventions that public health officials implement in every community, because there’s no single measure capable of counteracting the long-term effects of distrusts and hesitancy.[8,44,45]

This study has limitations. First, to preserve the consistency of the explanatory variables, we were limited in the diversity of information we included as potential predictors. While we believe all socioeconomic and demographic domains are well represented in the analysis, it would have been interesting to explore other religious, psychological, and occupational factors that have been associated with vaccine hesitancy.[11] Furthermore, it is possible that the specific status of the COVID-19 epidemic at the moment of the survey might have influenced the respondents’ perceptions which would have warranted the addition of the case rate or other epidemiological data. However, the differences in reporting and data-sharing agreements across states could have introduced bias to the estimates. Second, the nature of our hesitancy estimates and our methodological design, led us to quantify associations at a population level, which allowed us to better understand the drivers of vaccine hesitancy. However, because it is aggregated data, it was not possible for us to observe the potential heterogeneity that occurs at more granular levels. Third, it is plausible that people with greater levels of distrust towards public health activities at large would have been reluctant to answer the survey, in which case the sample might be lacking some of the more extreme sentiments. It is unclear, however, how prevalent this self-selection bias could have been and if it could have been enough to bias our results.

## Conclusions

To our knowledge, this is the first study that aims to identify the most important predictors of COVID-19 vaccine hesitancy at a population level in the United States. We found that economic living conditions such as income and poverty, marital status, race/ethnicity, and political party affiliation are the most important drivers of hesitancy. These results can help improve our understanding of the populations at higher risk of rejecting healthcare preventive measures. Therefore, they can be leveraged to better target interventions to increase the overall uptake of vaccines in future health emergencies.

## Data Availability

All data produced in the present study are available upon reasonable request to the authors.

